# Arbovirus surveillance in pregnant women in north-central Nigeria, 2019-2022

**DOI:** 10.1101/2023.08.04.23293671

**Authors:** Jerry Ogwuche, Charlotte Ajeong Chang, Olukemi Ige, Atiene S. Sagay, Beth Chaplin, Makshwar L. Kahansim, Michael Paul, Michael Elujoba, Godwin Imade, Georgenia Kweashi, Yu-Ching Dai, Szu-Chia Hsieh, Wei-Kung Wang, Donald J. Hamel, Phyllis J. Kanki

**Affiliations:** Our Lady of Apostles Hospital, Jos, Nigeria; Department of Immunology and Infectious Diseases, Harvard T.H. Chan School of Public Health, Boston, Massachusetts, USA; Jos University Teaching Hospital, University of Jos, Jos, Nigeria; Department of Tropical Medicine, Medical Microbiology and Pharmacology, John A. Burns School of Medicine, University of Hawaii at Manoa, Honolulu, Hawaii, USA

**Keywords:** Zika, dengue, chikungunya virus, arbovirus, pregnancy, microcephaly, West Africa, Nigeria

## Abstract

The adverse impact of Zika (ZIKV), dengue (DENV), and chikungunya (CHIKV) virus infection in pregnancy has been recognized in Latin America and Asia but is not well studied in Africa.

In Nigeria, we screened 1006 pregnant women for ZIKV, DENV and CHIKV IgM/IgG by rapid test (2019-2022). Women with acute infection were recruited for prospective study and infants were examined for any abnormalities from delivery through six months. A subset of rapid test-reactive samples were confirmed using virus-specific ELISAs and neutralization assays.

Prevalence of acute infection (IgM+) was 3.8%, 9.9% and 11.8% for ZIKV, DENV and CHIKV, respectively; co-infections represented 24.5% of all infections. Prevalence in asymptomatic women was twice the level of symptomatic infection. We found a significant association between acute maternal ZIKV/DENV/CHIKV infection and any gross abnormal birth outcome (p=0.014).

Further prospective studies will contribute to our understanding of the clinical significance of these endemic arboviruses in Africa.

## Introduction

In recent times, the emergence and re-emergence of Zika (ZIKV), dengue (DENV), and chikungunya (CHIKV) viruses have caused significant outbreaks of human disease globally (*1, 2*). Genomic and phylogenetic studies have delineated the origins and evolution of these arboviruses, which each have multiple Asian or American strains and lineages that expanded from ancestral African strains (*3, 4*). Although originally discovered in sub-Saharan Africa, research on these viruses and their biology in people on the continent has been limited. Arbovirus outbreaks in Africa likely go unrecognized due to their non-specific clinical presentations shared with malaria and typhoid and the lack of diagnostic capabilities for surveillance.

Across the globe, 90% of the estimated 200 million annual pregnancies occur in geographic regions with endemic or epidemic arboviruses (*5*). The 2015-2016 ZIKV outbreak in the Americas identified the teratogenic threat of ZIKV and renewed interest in the impact of other arboviruses on pregnancy and infant outcomes. Studies from outbreaks of ZIKV, DENV, and CHIKV have shown an increased risk of severe disease in pregnant women as well as fetal loss, prematurity, microcephaly and other birth defects or severe neonatal infection (*5*). While ZIKV, DENV and CHIKV infections may have similar clinical presentations, they exert temporally distinct impacts over the course of pregnancy. While antenatal ZIKV may result in microcephaly and congenital Zika syndrome (CZS), antenatal DENV is associated with preterm delivery and stillbirth, and CHIKV primarily impacts intra-partum outcomes resulting in neonatal disease and neurodevelopmental delays.

Nigeria is the most populous country in Africa, with over 200 million inhabitants (*6*), it is large (357,000 square miles) with diverse terrains and climatic regions, and over 70% of the population lives in rural areas. Global environmental mapping studies suggest that in Africa, roughly 453 million people live in areas suitable for ZIKV transmission, of which 111 million live in Nigeria, the nation at highest risk on the continent (*7*). Previously, we reported that ZIKV has been circulating in West Africa for at least two decades (*8*). The seroprevalence of ZIKV IgM was 6.4% in Nigerian patients and phylogenetic analysis showed that the ZIKVs belonged to the African lineage. As noted by the WHO, a linkage between a ZIKV strain circulating in Africa and microcephaly or other complications would have a significant impact on global risk assessments (*9*). The ZIKV outbreak in the Americas raised the question of whether pre-existing cross-reactive immunity or herd immunity may have resulted in the lack of detection of ZIKV-induced microcephaly in Africa (*10*).

In recent decades, DENV has emerged as a public health threat in Asia and Latin America. Although this arbovirus has been described in Africa since the late 19^th^ century and the *Aedes spp*. mosquito vector is ubiquitous, the epidemiology and public health importance in Africa is largely unknown (*11*). Although DENV outbreaks have been reported on the continent, diagnostics are lacking and febrile illnesses are frequently attributed to malaria. Halstead and others have considered genetic factors may be protective from disease based on the differential disease manifestations by race, as seen in outbreaks in Cuba (*12, 13*). Others have suggested that circulating endemic flaviviruses in Africa may provide cross-protection from severe dengue disease (*14*).

The impact of DENV infection during pregnancy on birth outcomes has been reported largely in case reports and case series from Asia and the Americas (*5*). In a meta-analysis, symptomatic dengue disease in pregnant women was associated with an increased risk of miscarriage (OR=3.51, 95% CI 1.15-10.77), stillbirth (RR=6.7, 95% CI 2.1-21.3), preterm birth (OR=1.71, 95% CI 1.06-2.76) and low birthweight for gestational age (1.41, 0.90-2.21) (*15*).

Chikungunya is an alphavirus transmitted, like ZIKV and DENV, by *Aedes aegypti* and *Aedes albopictus. Aedes albopictus* have been shown to deliver more than one arbovirus in their saliva (*16*). *Aedes aegypti* could be infected with and transmit all combinations of CHIKV, DENV-2 and ZIKV (*17, 18*). The different clinical consequences of single versus multiple co-infections are not clear (*19, 20*). The 2005-6 CHIKV outbreak on Reunion Island was estimated to have infected one-third of the African island’s population (>250,000) (*21*). The risk of CHIKV vertical transmission was highest in the intrapartum period at ∼50% transmission (*22*). Severe neonatal disease was also noted with sepsis-like illness, encephalitis, convulsions and death; exceptional cases of fetal demise were attributed to in utero CHIKV (*23*). In subsequent follow-up of the cohort of CHIKV-exposed children, 51% had neurodevelopmental delays compared to 15% of uninfected controls (*24, 25*).

Despite significant global risk for disease by ZIKV, DENV and CHIKV, our understanding of the epidemiology and pathogenesis of these arboviruses largely relies on research conducted in Asia and the Americas. The ZIKV epidemic in the Americas and its link to birth defects raised new research questions on the consequences of arbovirus infection in pregnancy. Given the high risk for arbovirus infection in Africa, our prospective study of symptomatic and asymptomatic pregnant women in Nigeria sought to better describe the epidemiology of arbovirus infection in West Africa and its potential impact on pregnancy and infant outcomes.

## Methods

### Study design and participants

Pregnant women ≥18 years of age attending the antenatal clinics at Jos University Teaching Hospital and Our Lady of Apostles Hospital in Jos, north-central Nigeria were recruited for arbovirus screening from April 1, 2019 to January 31, 2022. Women presenting with any of six common Zika symptoms: fever ≥37.5**°**C, rash, headache, arthralgia, conjunctivitis, or myalgia were recruited for a screening questionnaire and rapid test for ZIKV, DENV and CHIKV. One asymptomatic woman was recruited for every four symptomatic women. All women provided written informed consent for the screening, and separate informed consent for participation with their infants in the prospective follow-up study. The study was approved by the institutional review boards of the Jos University Teaching Hospital and the Harvard T.H. Chan School of Public Health.

Participating women provided a finger prick sample for the DPP**^®^**ZCD IgM/IgG rapid test (ChemBio, Medford, NY). They were administered a questionnaire that collected information on participant demographics, clinical history, exposure, and symptoms and had a brief clinical examination. The rapid test result was provided to the women within 30 minutes along with appropriate counseling. The DPP**^®^**ZCD IgM/IgG rapid test is CE-marked with 99-100% sensitivity and specificity for each virus and antibody isotype. All ZIKV, DENV, or CHIKV rapid test IgM and IgM/IgG positive women (symptomatic or asymptomatic) were recruited for the observational prospective study. At all routine antenatal visits and through delivery, women were examined for symptoms and provided blood and urine samples, a subset of which were tested for IgM antibodies by ELISA, neutralization tests and for nucleic acid detection.

At delivery, infants were examined for any abnormal clinical outcomes, including microcephaly and signs of congenital Zika syndrome. Any adverse pregnancy outcomes, including miscarriage, stillbirth, and premature delivery were also documented. Infants were followed at their routine follow-up visits: 6 weeks, 10 weeks, 14 weeks, and 6 months. They were observed for any developmental abnormalities, including ocular examinations. Microcephaly was defined as head circumference with Z-score less than or equal to −2 standard deviations (SD), calculated as the difference between the head circumference and the median head circumference of a reference population (same age and sex) divided by the SD for the reference population used by the WHO Multicentre Growth Reference Study (*26*). An infant was defined as low weight if their birth weight Z-score was less than −2 SD for their age and sex. An infant was defined as preterm if delivered earlier than 37 weeks gestational age, the Fenton growth charts were used to calculate Z-scores for head circumference and weight for preterm infants, as used at the hospitals (*27, 28*).

### Confirmation of arbovirus serology

Anti-ZIKV and anti-DENV IgM and IgG were determined by previously described ELISAs using DENV and ZIKV envelope protein fusion-loop (FL) mutated virus-like particles (VLP) as antigens, which greatly reduced cross-reactivities from other flaviviruses (*29*). The sensitivity/specificity of DENV FL-VLP-based IgM and IgG ELISAs were 75.0/95.8% and 100.0/93.0%, respectively; the sensitivity/specificity of ZIKV FL-VLP-based IgM and IgG ELISAs were 90.0/99.3% and 100.0/83.3%, respectively. Previously described ZIKV and DENV NS1 IgG ELISAs were also used to distinguish previous DENV and ZIKV infections (*30*).

Microneutralization test using Vero cells and DENV (DENV1-Hawaii, DENV2-NGC, DENV3-H87, DENV4-H241), or ZIKV (PRVABC59 strain) was performed in 96-well plates as previously described, and 90% neutralization (NT90) titers to DENV1-4 and ZIKV were determined (*31*). NT90 titers <10 to all 5 flaviviruses tested were designated as DENV- and ZIKV-naïve, ≥10 to only one virus as primary infection (*32*), ≥10 to two or more viruses as multiple flavivirus infections. The neutralization test for CHIKV was performed using a CHIKV pseudovirus assay (*33*). The % neutralization was determined and NT50 titer was the serum dilution that reached 50% neutralization using 4-parameter nonlinear regression analysis (GraphPad 6.0).

We analyzed the sensitivity and specificity of the DPP**^®^**ZCD IgM/IgG rapid test using 146 convalescent-phase samples from RT-PCR-confirmed DENV, ZIKV and CHIKV cases, and naïve samples (*33–35*). The sensitivity/specificity of CHIKV-, ZIKV- and DENV-IgM was 90.9/95.1%, 81.0/94.2% and 80.0/82.0%, respectively (Supplementary Table 1).

Viral RNA was subjected to nested or semi-nested PCR targeting a conserved region of the NSP2 gene of CHIKV, 3’-untranslated region of DENV, and NS5 or NS4B gene of ZIKV (*36, 37*). Electrophoresis of RT-PCR products and bands with the predicted size were purified for sequencing; assay details provided upon request.

### Statistical analysis

Descriptive statistics of demographic and clinical data described the study population. The Χ^2^ test was used in bivariate analyses to test for associations between demographic and exposure variables and arbovirus infection. Variables significant at p≤0.20 in bivariate analyses were included in the multivariate logistic regression model, and backwards elimination used to build the final model predicting acute arbovirus infection, which retained variables significant at p≤0.05.

For the prospective follow-up study, we used the two-sided Fisher’s exact test to compare outcomes of infants born to arbovirus rapid test IgM-positive mothers to outcomes recorded in the general labor and delivery population registers at JUTH and OLA; we repeated this analysis with just the infants born to ELISA- and NT-confirmed arbovirus-positive mothers. For the overall comparison of any abnormal birth outcome, study infants were not double-counted if they had more than one abnormal category. Stata v.15.1 (StataCorp, College Station, TX, USA) software was used for the statistical analyses.

## Results

From April 2019 to January 2022, we recruited 1006 consenting pregnant women at their antenatal care visit, 478 at JUTH and 528 at OLA. One-third of women (312/1006, 31.0%) showed antibody responses to one or more of the arboviruses, with 19.9% IgM or IgM/IgG reactive and 11.1% reactive to IgG only (Figure 1). Acute or recent arbovirus prevalence, defined by IgM or IgM/IgG seropositivity, was ZIKV 1.4% (14/1006), DENV 6.0% (60/1006) and CHIKV 7.7% (77/1006), ZIKV+DENV 0.7% (7/1006), ZIKV+CHIKV 0.9% (9/1006), DENV+CHIKV 2.5% (25/1006) and ZIKV+CHIKV+DENV 0.8% (8/1006) (Table 1).

**Figure 1:**
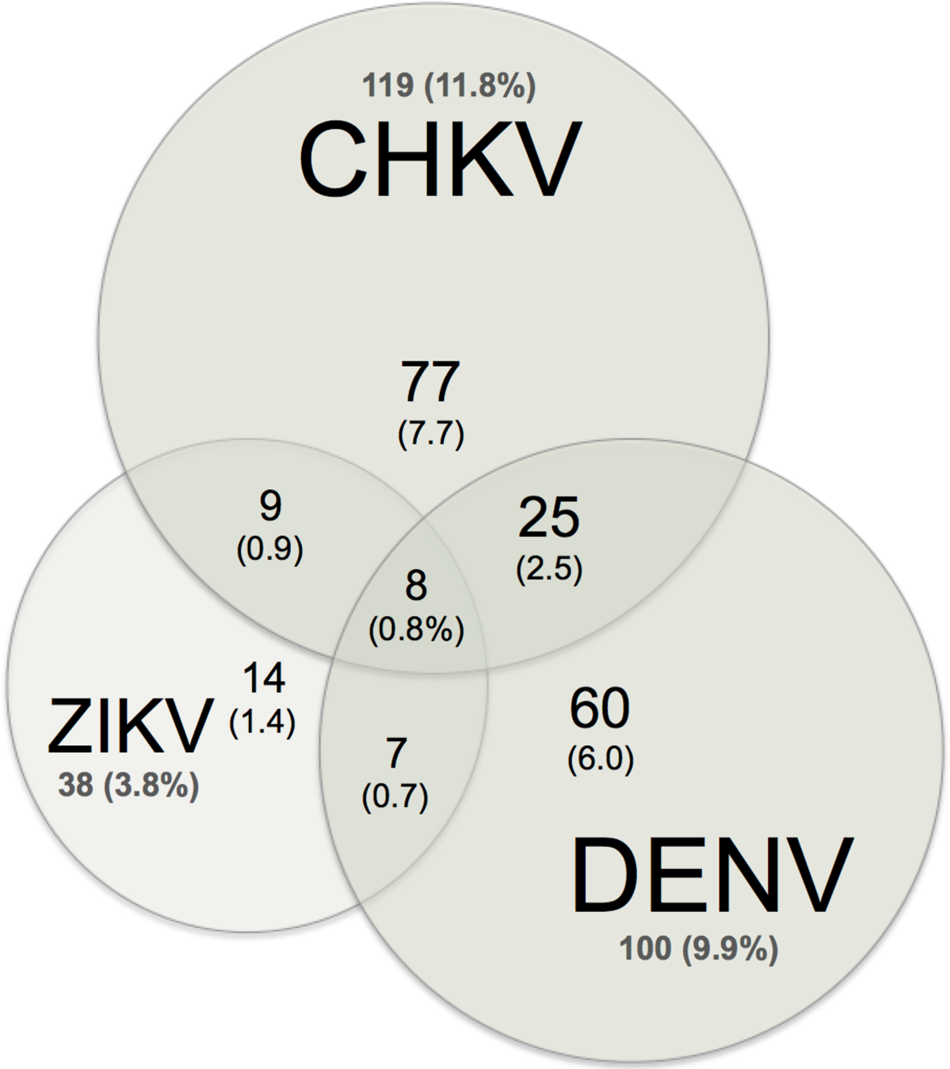
Venn diagram of acute (IgM) ZIKV, DENV and CHIKV infection Women with any IgM reactivity (200, 19.9%) by arbovirus.

**Table 1.** Prevalence of Zika, chikungunya, and dengue infection (Chembio IgM+) among pregnant women at Jos University Teaching Hospital and Our Lady of Apostles Hospital, Jos, Nigeria, April 2019–January 2022*.

|  | Any IgM+ (with or without IgG+) |  |  | IgG+-only |  |  |
| --- | --- | --- | --- | --- | --- | --- |
|  | Symptomatic, no. (%) | Asymptomatic, no. (%) | Total, no. (%) | Symptomatic, no. (%) | Asymptomatic, no. (%) | Total, no. (%) |
| Total | 787 | 219 | 1006 | 787 | 219 | 1006 |
| ZIKV | 10 (1.3%) | 4 (1.8%) | 14 (1.4%) | 4 (0.5%) | 0 (0.0%) | 4 (0.4%) |
| CHIKV | 53 (6.7%) | 24 (11.0%) | 77 (7.7%) | 38 (4.8%) | 12 (5.5%) | 50 (5.0%) |
| DENV | 39 (5.0%) | 21 (9.6%) | 60 (6.0%) | 26 (3.3%) | 5 (2.3%) | 31 (3.1%) |
| ZIKV & CHIKV | 6 (0.8%) | 3 (1.4%) | 9 (0.9%) | 3 (0.4%) | 0 (0.0%) | 3 (0.3%) |
| ZIKV & DENV | 5 (0.6%) | 2 (0.9%) | 7 (0.7%) | 5 (0.6%) | 3 (1.4%) | 8 (0.8%) |
| CHIKV & DENV | 12 (1.5%) | 13 (5.9%) | 25 (2.5%) | 10 (1.3%) | 4 (1.8%) | 14 (1.4%) |
| ZIKV, CHIKV, & DENV | 7 (0.9%) | 1 (0.5%) | 8 (0.8%) | 2 (0.3%) | 0 (0.0%) | 2 (0.2%) |
| Arbovirus reactive | 132 (16.8%) | 68 (31.1%) | 200 (19.9%) | 88 (11.2%) | 24 (11.0%) | 112 (11.1%) |

\*IgM, immunoglobulin M; IgG, immunoglobulin G; ZIKV, Zika virus; CHIKV, chikungunya virus; DENV, dengue virus.

Acute infected women with IgM+ reactivity showed significant variation by arbovirus and time. Higher recruitment of symptomatic women corresponded to the three rainy seasons over the study period (Figure 2). To evaluate temporal trends, the study was divided into three time periods: April 2019-March 2020, April 2020-March 2021, and April 2021-January 2022. The IgM+ prevalence for any arbovirus was 26.7% (88/330) in the first time period and reduced to 16.0% (59/368) in period 2 and 17.2% (53/308) in period 3. Age-specific arbovirus prevalence demonstrated a characteristic pattern of endemic infection (Supplementary Figure S1).

**Figure 2:**
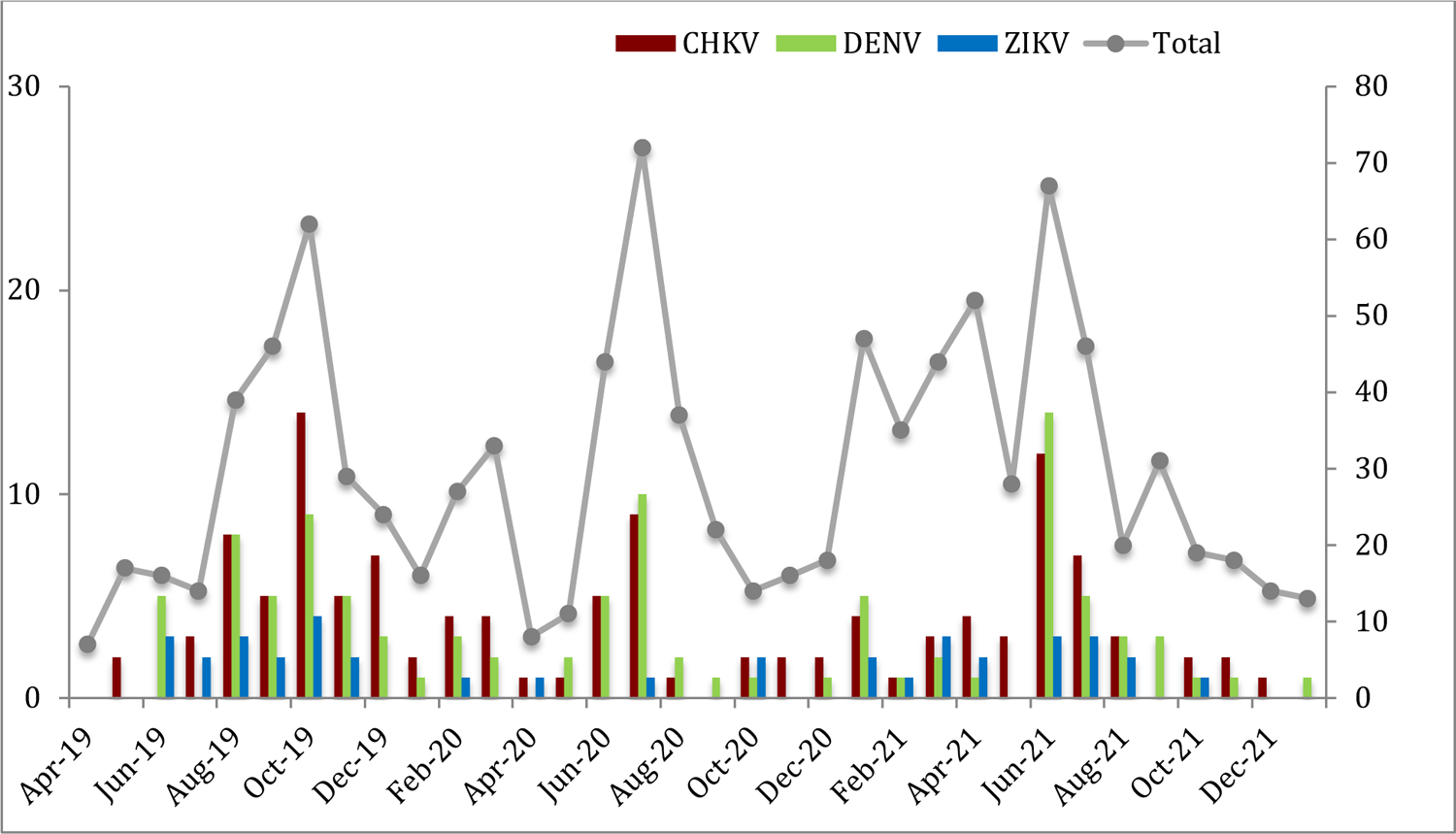
Acute (IgM) arbovirus infection (2019-2022), Seroprevalence (left axis) and number of women tested (right axis); rainy season indicated by rain cloud.

We analyzed baseline characteristics of pregnant women to identify risks for acute (IgM) arbovirus infection (Table 2). In bivariate analysis, OLA clinic, study year 2019-2020, age 25-34 years, Igbo/other ethnicity, and unemployment status were associated with acute infection.

**Table 2.**
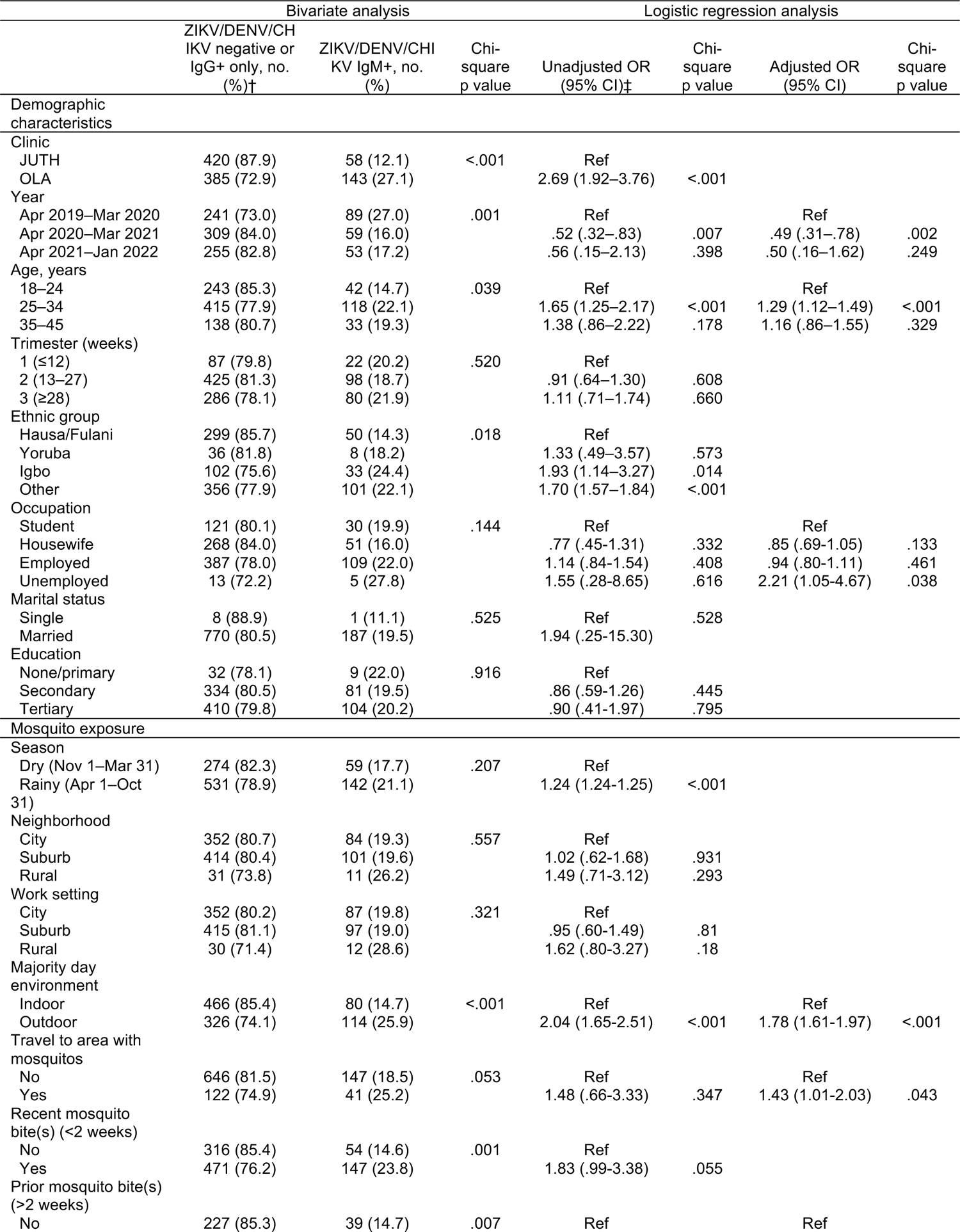
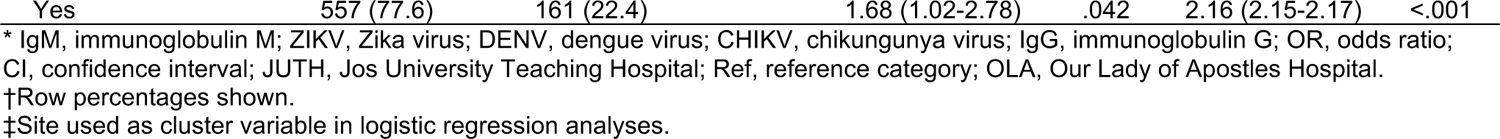
Baseline characteristics and risk factors for Zika, dengue, and chikungunya infection (Chembio IgM+) among pregnant women in Jos, Nigeria, April 2019-January 2022*.

|  | Bivariate analysis |  |  | Logistic regression analysis |  |  |  |
| --- | --- | --- | --- | --- | --- | --- | --- |
|  | ZIKV/DENV/CHIKV negative or IgG+ only, no. (%)† | ZIKV/DENV/CHIKV IgM+, no. (%) | Chi-square p value | Unadjusted OR (95% CI)‡ | Chi-square p value | Adjusted OR (95% CI) | Chi-square p value |
| Demographic characteristics |  |  |  |  |  |  |  |
| Clinic |  |  |  |  |  |  |  |
| JUTH | 420 (87.9) | 58 (12.1) | <.001 | Ref |  |  |  |
| OLA | 385 (72.9) | 143 (27.1) |  | 2.69 (1.92–3.76) | <.001 |  |  |
| Year |  |  |  |  |  |  |  |
| Apr 2019–Mar 2020 | 241 (73.0) | 89 (27.0) | .001 | Ref |  | Ref |  |
| Apr 2020–Mar 2021 | 309 (84.0) | 59 (16.0) |  | .52 (.32–.83) | .007 | .49 (.31–.78) | .002 |
| Apr 2021–Jan 2022 | 255 (82.8) | 53 (17.2) |  | .56 (.15–2.13) | .398 | .50 (.16–1.62) | .249 |
| Age, years |  |  |  |  |  |  |  |
| 18–24 | 243 (85.3) | 42 (14.7) | .039 | Ref |  | Ref |  |
| 25–34 | 415 (77.9) | 118 (22.1) |  | 1.65 (1.25–2.17) | <.001 | 1.29 (1.12–1.49) | <.001 |
| 35–45 | 138 (80.7) | 33 (19.3) |  | 1.38 (.86–2.22) | .178 | 1.16 (.86–1.55) | .329 |
| Trimester (weeks) |  |  |  |  |  |  |  |
| 1 (≤12) | 87 (79.8) | 22 (20.2) | .520 | Ref |  |  |  |
| 2 (13–27) | 425 (81.3) | 98 (18.7) |  | .91 (.64–1.30) | .608 |  |  |
| 3 (≥28) | 286 (78.1) | 80 (21.9) |  | 1.11 (.71–1.74) | .660 |  |  |
| Ethnic group |  |  |  |  |  |  |  |
| Hausa/Fulani | 299 (85.7) | 50 (14.3) | .018 | Ref |  |  |  |
| Yoruba | 36 (81.8) | 8 (18.2) |  | 1.33 (.49–3.57) | .573 |  |  |
| Igbo | 102 (75.6) | 33 (24.4) |  | 1.93 (1.14–3.27) | .014 |  |  |
| Other | 356 (77.9) | 101 (22.1) |  | 1.70 (1.57–1.84) | <.001 |  |  |
| Occupation |  |  |  |  |  |  |  |
| Student | 121 (80.1) | 30 (19.9) | .144 | Ref |  | Ref |  |
| Housewife | 268 (84.0) | 51 (16.0) |  | .77 (.45–1.31) | .332 | .85 (.69–1.05) | .133 |
| Employed | 387 (78.0) | 109 (22.0) |  | 1.14 (.84–1.54) | .408 | .94 (.80–1.11) | .461 |
| Unemployed | 13 (72.2) | 5 (27.8) |  | 1.55 (.28–8.65) | .616 | 2.21 (1.05–4.67) | .038 |
| Marital status |  |  |  |  |  |  |  |
| Single | 8 (88.9) | 1 (11.1) | .525 | Ref | .528 |  |  |
| Married | 770 (80.5) | 187 (19.5) |  | 1.94 (.25–15.30) |  |  |  |
| Education |  |  |  |  |  |  |  |
| None/primary | 32 (78.1) | 9 (22.0) | .916 | Ref |  |  |  |
| Secondary | 334 (80.5) | 81 (19.5) |  | .86 (.59–1.26) | .445 |  |  |
| Tertiary | 410 (79.8) | 104 (20.2) |  | .90 (.41–1.97) | .795 |  |  |
| Mosquito exposure |  |  |  |  |  |  |  |
| Season |  |  |  |  |  |  |  |
| Dry (Nov 1–Mar 31) | 274 (82.3) | 59 (17.7) | .207 | Ref |  |  |  |
| Rainy (Apr 1–Oct 31) | 531 (78.9) | 142 (21.1) |  | 1.24 (1.24–1.25) | <.001 |  |  |
| Neighborhood |  |  |  |  |  |  |  |
| City | 352 (80.7) | 84 (19.3) | .557 | Ref |  |  |  |
| Suburb | 414 (80.4) | 101 (19.6) |  | 1.02 (.62–1.68) | .931 |  |  |
| Rural | 31 (73.8) | 11 (26.2) |  | 1.49 (.71–3.12) | .293 |  |  |
| Work setting |  |  |  |  |  |  |  |
| City | 352 (80.2) | 87 (19.8) | .321 | Ref |  |  |  |
| Suburb | 415 (81.1) | 97 (19.0) |  | .95 (.60–1.49) | .81 |  |  |
| Rural | 30 (71.4) | 12 (28.6) |  | 1.62 (.80–3.27) | .18 |  |  |
| Majority day environment |  |  |  |  |  |  |  |
| Indoor | 466 (85.4) | 80 (14.7) | <.001 | Ref |  | Ref |  |
| Outdoor | 326 (74.1) | 114 (25.9) |  | 2.04 (1.65–2.51) | <.001 | 1.78 (1.61–1.97) | <.001 |
| Travel to area with mosquitos |  |  |  |  |  |  |  |
| No | 646 (81.5) | 147 (18.5) | .053 | Ref |  | Ref |  |
| Yes | 122 (74.9) | 41 (25.2) |  | 1.48 (.66–3.33) | .347 | 1.43 (1.01–2.03) | .043 |
| Recent mosquito bite(s) (<2 weeks) |  |  |  |  |  |  |  |
| No | 316 (85.4) | 54 (14.6) | .001 | Ref |  |  |  |
| Yes | 471 (76.2) | 147 (23.8) |  | 1.83 (.99–3.38) | .055 |  |  |
| Prior mosquito bite(s) (>2 weeks) |  |  |  |  |  |  |  |
| No | 227 (85.3) | 39 (14.7) | .007 | Ref |  | Ref |  |

\* IgM, immunoglobulin M; ZIKV, Zika virus; DENV, dengue virus; CHIKV, chikungunya virus; IgG, immunoglobulin G; OR, odds ratio; CI, confidence interval; JUTH, Jos University Teaching Hospital; Ref, reference category; OLA, Our Lady of Apostles Hospital.
†Row percentages shown.
‡Site used as cluster variable in logistic regression analyses.

Questionnaire data on mosquito exposure also found that majority of daytime outdoors, travel to locales with mosquitoes, and mosquito bites during pregnancy were associated with acute arbovirus infection. In multiple logistic regression using clinic as a cluster variable, age 25-34 years, study year 2019-2020, unemployment status, majority outdoor day environment, travel to locales with mosquitoes, and prior mosquito bites (>2 weeks) remained statistically significant.

Unexpectedly, lower rates of acute arbovirus infection were observed in women who reported symptoms at screening 132/787 (16.8%) compared to 68/219 (31.1%) in asymptomatic women (Fishers exact p<0.001), ratio of asymptomatic to symptomatic of 2:1. We failed to observe high levels of IgG seroprevalence or hyper-endemic arbovirus infection in the absence of IgM, with relatively low levels of IgG reactivity to all three arboviruses (1.7-6.9%).

We compared birth outcomes for 101 ZIKV/DENV/CHIKV rapid test IgM-positive women to 5930 JUTH and OLA delivery records from May 2019 to May 2021(Table 3). We found potential associations between maternal ZIKV/DENV/CHIKV infection during pregnancy and macerated stillbirth (p=0.017), asymmetric intrauterine growth retardation (p=0.017), microcephaly (p<0.001), cleft lip/palate (p=0.017), polydactyly (p=0.017), and multiple congenital anomalies (p<0.001) of the infant. We found a significant association between maternal ZIKV/DENV/CHIKV infection during pregnancy and any adverse or abnormal gross birth outcome (p=0.014).

**Table 3.** Abnormal delivery outcomes among study infants born to mothers with any acute Zika, dengue, or chikungunya infection versus infants in birth registers at Jos University Teaching Hospital and Our Lady of Apostles Hospital, Jos, Nigeria, May 2019–May 2021*†.

|  | Birth registers, no.<br>(%)‡ | Study infants born to<br>any Chembio<br>ZIKV/DENV/CHIKV<br>IgM+ mother, no.<br>(%) | Fisher's exact<br>p value | Study infants born to<br>any<br>ZIKV/DENV/CHIKV<br>IgM-confirmed<br>mother, no. (%) | Fisher's exact<br>p value |
| --- | --- | --- | --- | --- | --- |
| Total | 5930 | 101 |  | 37 |  |
| Macerated stillbirth | 117 (2.0%) | 6 (5.9%) | .017 | 3 (8.1%) | .038 |
| Fresh stillbirth | 81 (1.4%) | 0 (0.0%) | .647 | 0 (0.0%) | 1 |
| Preterm | 294 (5.0%) | 6 (5.9%) | .640 | 4 (10.8%) | .111 |
| Asymmetric intrauterine<br>growth retardation | 0 (0.0%) | 1 (1.0%) | .017 | 0 (0.0%) | ... |
| Neural tube defects | 7 (0.1%) | 0 (0.0%) | 1 | 0 (0.0%) | 1 |
| *Anencephalocele*§ | 3 (0.1%) | 0 (0.0%) | 1 | 0 (0.0%) | 1 |
| *Spina bifida* | 2 (0.0%) | 0 (0.0%) | 1 | 0 (0.0%) | 1 |
| *Encephalocele* | 1 (0.0%) | 0 (0.0%) | 1 | 0 (0.0%) | 1 |
| *Holoprosencephaly* | 1 (0.0%) | 0 (0.0%) | 1 | 0 (0.0%) | 1 |
| Other CNS abnormalities | 34 (0.6%) | 6 (5.9%) | <.001 | 2 (5.4%) | .021 |
| *Microcephaly* | 32 (0.5%) | 6 (5.9%) | <.001 | 2 (5.4%) | .019 |
| *Hydrocephalus* | 2 (0.0%) | 0 (0.0%) | 1 | 0 (0.0%) | 1 |
| Cleft lip/palate | 0 (0.0%) | 1 (1.0%) | .017 | 1 (2.7%) | .006 |
| Gastrointestinal defects | 4 (0.1%) | 0 (0.0%) | 1 | 0 (0.0%) | 1 |
| *Omphalocele* | 2 (0.0%) | 0 (0.0%) | 1 | 0 (0.0%) | 1 |
| *Duodenal atresia* | 2 (0.0%) | 0 (0.0%) | 1 | 0 (0.0%) | 1 |
| Limb defects | 1 (0.0%) | 1 (1.0%) | .033 | 1 (2.7%) | .012 |
| *Clubbed foot* | 1 (0.0%) | 0 (0.0%) | 1 | 0 (0.0%) | 1 |
| *Polydactyly* | 0 (0.0%) | 1 (1.0%) | .017 | 1 (2.7%) | .006 |
| Multiple gross congenital<br>anomalies¶ | 3 (0.1%) | 3 (3.0%) | <.001 | 2 (5.4%) | <.001 |
| Any abnormality# | 541 (9.1%) | 17 (16.8%) | .014 | 9 (24.3%) | .005 |

\*ZIKV, Zika virus; DENV, dengue virus; CHIKV, chikungunya virus; IgM, immunoglobulin M.
†Data shown includes adverse and gross abnormal outcomes recorded in hospital birth registers; other outcomes, such as neonatal sepsis and jaundice, are not shown. 101 study infants born to mothers with a Chembio IgM+ rapid test result during the time period were observed at birth. Among them, 37 of the mothers had their IgM+ status confirmed by ELISA or neutralization assay.
‡Column percentages shown.
§Italicized outcomes are subsets of their larger category. Numbers, percentages, and p values are shown for each outcome and category, if applicable.
¶Among the three study infants with multiple congenital anomalies, one also had cleft lip/palate, asymmetric microcephaly, craniocynostosis, hypotonia, and died on day 2; another also had loss of consciousness, seizures, hypotonia, and sustained microcephaly through 6 months; another was a stillbirth with facial dysmorphism.

We conducted additional confirmatory serologic studies on a subset of samples from the arbovirus IgM-positive women. Using FL-mutated VLP ELISAs, NS1-based ELISAs, microneutralization or pseudovirus neutralization tests, we confirmed 37 samples and re-compared the delivery outcomes of these confirmed arbovirus infected women to the overall hospital control deliveries (Table 3). Microcephaly was associated with confirmed maternal IgM-positive status (p=0.019), confirmed maternal ZIKV/DENV/CHIKV infection during pregnancy remained significantly associated with any gross abnormal birth outcome (p=.005). We also identified and described 33 infants from May 2019-May 2021 with any abnormal observations recorded from birth through six months, including transient weight or head circumference and neonatal jaundice or sepsis, which were not recorded in the birth registers, with the mother’s serology results (Supplementary Table S2).

## Discussion

Our surveillance study in pregnant women provides new insights into the epidemiology of arbovirus infection in a West African setting. Spanning three rainy seasons, our study detected regular peaks in symptomatic pregnant women enrollment coincident with rainy seasons and heightened risk for mosquito transmission. We described acute infection with ZIKV, DENV and CHIKV continuously throughout the 34-month study, which decreased during the COVID-19 pandemic, with an overall IgM prevalence of 3.8% ZIKV, 9.9% DENV, and 11.8% CHIKV.

Geographic regions with endemic *Aedes sp*. may be at risk for co-circulation of ZIKV, DENV and CHIKV, as well as other related and serologically cross-reactive viruses such as Yellow Fever virus (YFV), West Nile virus (WNV), Usutu virus and the alphavirus O’nyong-nyong virus. The similar non-distinguishing and transient clinical presentations and diagnostic assays compromised by cross-reactivity hinder disease surveillance efforts. Comprehensive arbovirus surveillance is rarely performed in most African settings where inadequate infrastructure and/or the cost of testing are prohibitive. As a result, most studies are cross-sectional surveys on febrile patients that focus on a single viral pathogen, limiting our understanding of arbovirus co-infection. We found significant ZIKV, DENV, and CHIKV co-infections in pregnant women comprising 24.5% of all IgM-positive infections. Co-infection with CHIKV and DENV (with or without ZIKV) represented 67.3% of the co-infections and 18.4% ZIKV-CHIKV. In Brazil and Colombia, pregnant women with multiple arbovirus infections have been described (*38, 39*). Case reports of congenital co-infection of CHIKV and ZIKV described increased disease manifestations often leading to miscarriage (*40*). The clinical significance of single versus multiple co-infections remains unclear.

Although DENV, ZIKV and CHIKV have historical origins in Africa, severe arbovirus disease outbreaks have been only rarely described. For ZIKV, the rapid geographic dispersal and distinct disease manifestations in the 2015-16 epidemic in the Americas and the re-emergence in the 2015 Cape Verde and 2016 Guinea Bissau outbreaks on the African continent suggest that this may be changing (*9*). However, it is notable that these two outbreaks were associated with the ZIKV of Asian lineage. We have previously reported a 6% seroprevalence in febrile West Africans (2004–2016) with African lineage ZIKV (*8*), similar to the reported 6.2% IgM seroprevalence by ZIKV NS1-based ELISA in north-central Nigeria in 2018 (*41*). This low prevalence profile may be explained by the mosquito vectors; lower ZIKV transmission potential has been described for native *A. aegpyti* species compared to sub-species from Asia or the Americas, also seen in YFV and DENV transmission (*42*).

Worldwide, DENV is considered one of the most widely distributed arboviruses with over 100 million annual infections. An analysis of 2020 data from 23 countries in southeast Asia and Latin America described reduced dengue transmission associated with COVID-19 restrictions which could not be explained by seasonal dengue cycles or underreporting (*43, 44*). In our time period analysis, we also saw a reduction in acute IgM-positive arbovirus infections in pregnant women from 26.7% in the first time period to 16.0% and 17.2% from 2020-2022, coinciding with the COVID-19 pandemic. The Nigerian governmental response to the COVID-19 pandemic began in March 2020, which closed national and state borders and instituted weeks of stay-at-home orders, followed by restricted movement and curfews. Antenatal attendance dropped by 50% for several months and COVID-19 surges along with other imposed public health restrictions likely limited population movement and gathering.

In the literature, most studies of ZIKV and DENV are based on hospital-based disease surveillance whereby patients presenting with fever or other significant flavivirus symptoms present to healthcare settings for care. By contrast, our study population consisted of healthy and mildly symptomatic pregnant women attending antenatal clinics (Supplementary Table S3). We found that the rate of acute arbovirus infection was two times higher in asymptomatic women (31.1%) compared with mildly symptomatic women (16.8%). Other arbovirus studies have reported that most infections are asymptomatic and symptoms non-specific, with the ratio of symptomatic to inapparent infection ranging from 1:1 to 1:19 with DENV and 1:0.03 to 1:1.2 with CHIKV (*45–48*). In the 2007 ZIKV outbreaks in Yap Island, household surveillance estimated a ratio of 1 symptomatic to every 4.4 asymptomatic infected individuals (*49, 50*). Endemic exposure in Africa and Asia may limit recognized outbreaks via herd immunity, which could be imparted by multiple cross-reacting flaviviruses.

High arbovirus seroprevalence rates have been reported in serosurveys among febrile hospital outpatients in Nigeria. In a meta-analysis of CHIKV in Nigeria, 7 cross-sectional studies contributed to the pooled anti-CHIKV IgM prevalence of 26.7% and 29.3%; these studies were conducted in the North-east and South-west of the country, where geographic region was a predictor for high seroprevalence (*51*). In contrast, our study demonstrated lower rates of both acute IgM and convalescent/remote IgG seroreactivity throughout the study period, consistent with our previous studies (*8*). Differences in geographic region, health status of study participants and serologic assays employed may provide some explanation for the apparent differences in results from our study and previous studies in the country.

Microcephaly in ZIKV infected infants in the 2015-16 outbreak of the Americas increased interest in the pathologic effects of these previously poorly studied flaviviruses. ZIKV, DENV and CHIKV have been associated with adverse pregnancy outcomes including miscarriage, stillbirths and perinatal death (*22, 52, 53*). In our study, microcephaly was found in 6/101 (5.9%) acute arbovirus-infected women compared to 32/5930 (0.5%) in the control hospital deliveries, (p<0.001). We were not able to confirm arbovirus transmission in all infants born to IgM+ mothers, so we are cautious in interpreting these results. Nonetheless, the significant differences in infants born to arbovirus-infected women compared to the community control population warrant future study. Acute arbovirus infection was significantly associated with adverse and gross abnormal birth outcomes in both rapid test and confirmed subset analysis (p=0.014 and 0.005, respectively). We also report significant associations with multiple congenital anomalies (p<0.001), which should be studied further.

Our study has several limitations. The low-level prevalence rates of individual arbovirus infections and high rates of co-infections made direct associations with maternal and infant outcomes difficult. We were not able to confirm all rapid test IgM/IgG samples with other confirmatory assays. As our study population was pregnant women, some arbovirus symptoms are conflated with common pregnancy symptoms. The prospective study design and evaluation of multiple arboviruses was a strength of our study. We were able to describe acute and convalescent-phase arbovirus infection, frequent co-infection, and significant acute arbovirus infection in asymptomatic pregnant women. Most importantly, with evaluation of ∼6000 deliveries at our study clinics we describe a significant association of birth abnormalities in pregnant women with acute arbovirus infection. Larger studies are needed to verify these associations and identify risk factors for infant abnormalities given arboviral infection/coinfection during pregnancy.

ZIKV, DENV and CHIKV are arboviruses of global public health importance. While their dynamic epidemiology and association with clinical disease are actively studied in the Americas and Asia, the biological consequences of these endemic viruses in Africa remain poorly understood. Our study demonstrated low-level but persistent risk and exposure of all three viruses in pregnant women in Nigeria. The significant rates of asymptomatic infection and potential association with abnormal birth outcomes warrant further epidemiologic research to reveal the burden of disease, risks of transmission, and immunopathogenesis linked to disease outcomes of these viruses on the continent.

## Data Availability

All data produced in the present study are available upon reasonable request to the authors.

## Acknowledgments

We acknowledge the invaluable support of our clinic and laboratory staff at JUTH and OLA that enabled this study.

