## Supplementary Tables 1, 2 and 3 for "Arbovirus surveillance in pregnant women in north-central Nigeria, 2019-2022"

Supplementary Table S1: Results of DPP® ZCD IgM/IgG rapid test among convalescent-phase samples from RT-PCR-confirmed ZIKV, DENV and CHIKV cases

| DPP® ZCD IgM/IgG rapid test | No. of positive/total samples (%) in different serum/plasma panels* | | | | | |
|---|---|---|---|---|---|---|
| | naïve | pDENV | sDENV | pZIKV | ZIKV wprDENV | CHIKV |
| CHIKV IgM† | 2/47 (4.3%) | 1/11 (9.1%) | 1/24 (4.2%) | 4/21 (19.0%) | 6/21 (28.6%) | **20/22 (90.9%)** |
| ZIKV IgM† | 1/47 (2.1%) | 0/11 (0%) | 3/24 (12.5%) | **19/21 (90.5%)** | **15/21 (71.4%)** | 2/22 (9.1%) |
| DENV IgM† | 1/47 (2.1%) | **10/11 (90.9%)** | **18/24 (75.0%)** | 11/21 (52.4%) | 7/21 (33.3%) | 1/22 (4.5%) |

*pDENV, primary DENV infection; sDENV, secondary DENV infection; pZIKV, primary ZIKV infection; ZIKVwprDENV, ZIKV infection with previous DENV infection; CHIKV, CHIKV infection. Convalescent-phase serum/plasma samples were from RT-PCR-confirmed ZIKV, DENV and CHIKV cases as described previously (33-35).

†The overall sensitivity/specificity of CHIKV-, ZIKV- and DENV-IgM was 90.9/88.7%, 81.0/94.2% and 80.0/82.0%, respectively. The specificity of CHIKV-IgM was 95.1%, when the pZIKV and ZIKVwprDENV panels with unknown history of CHIKV infection were not included in the calculation.

Supplementary Table S2. Abnormal outcomes of study infants born to mothers with acute Zika, dengue, or chikungunya infection (Chembio IgM+) at Jos University Teaching Hospital and Our Lady of Apostles Hospital, Jos, Nigeria, May 2019–May 2021*‡

| Infant ID | Site | Mother's Chembio IgM reactivity | Delivery year | Infant sex | Infant outcome§ |
|---|---|---|---|---|---|
| 1 | JUTH | C | 2019 | Male | Sepsis |
| 2 | JUTH | C | 2019 | Male | Transient weight (normal>low) |
| 3 | JUTH | D | 2019 | Male | Preterm (36 weeks), skin lesions on face and neck |
| 4 | JUTH | C | 2019 | Female | Preterm (33 weeks), macerated stillbirth |
| 5 | JUTH | Z,D | 2020 | Female | Jaundice |
| 6 | JUTH | C | 2020 | Female | Sepsis |
| 7 | JUTH | Z,D,C | 2020 | Female | Jaundice |
| 8 | JUTH | D | 2020 | Female | Transient weight (low>normal), asymmetrical intrauterine growth retardation |
| 9 | JUTH | D | 2020 | Male | Skin rash |
| 10 | JUTH | Z,D,C | 2020 | Male | Macerated stillbirth |
| 11 | JUTH | C | 2021 | Female | Preterm (30 weeks) |
| 12 | OLA | C | 2019 | Female | Preterm (35 weeks), macerated stillbirth. Mother died in childbirth. |
| 13 | OLA | D | 2019 | Female | Preterm (35 weeks), transient weight (normal>low) transient head circumference (normal>small) |
| 14 | OLA | D,C | 2019 | Male | Plethora, macrosomia |
| 15 | OLA | D,C | 2019 | Male | Transient weight (normal>low) |
| 16 | OLA | C | 2019 | Female | Transient head circumference (small>normal) |
| 17 | OLA | C | 2019 | Female | Preterm (36 weeks) |
| 18 | OLA | C | 2019 | Female | Low weight, multiple congenital anomalies, including microcephaly (asymmetric), cleft lip/palate, distended abdomen, craniocynostosis, hypotonia, apnoeic attacks. Infant died day 2. |
| 19 | OLA | C | 2019 | Female | Jaundice, sepsis |
| 20 | OLA | Z | 2019 | Male | Transient head circumference (small>normal) |
| 21 | OLA | Z,D,C | 2019 | Female | Jaundice, sepsis, fever, skin lesions on neck |
| 22 | OLA | C | 2019 | Female | Jaundice, sepsis, anemia |
| 23 | OLA | D,C | 2019 | Male | Multiple congenital anomalies, including microcephaly, unconscious, hypotonia, seizures |
| 24 | OLA | D,C | 2019 | Female | Transient head circumference (small>normal) |
| 25 | OLA | Z | 2020 | Female | Transient head circumference (normal>small) |
| 26 | OLA | C | 2020 | Female | Transient head circumference (normal>small) |
| 27 | OLA | D | 2020 | Male | Macerated stillbirth, low weight |
| 28 | OLA | C | 2020 | Male | Polydactyly, jaundice, sepsis, fever |
| 29 | OLA | C | 2020 | Male | Transient weight (normal>low) |
| 30 | OLA | C | 2020 | Male | Macerated stillbirth, multiple congenital anomalies, including facial dysmorphism |
| 31 | OLA | C | 2020 | Female | Macerated stillbirth, microcephaly |
| 32 | OLA | Z | 2021 | Female | Transient weight (normal>low), sepsis, fever |
| 33 | OLA | C | 2021 | Male | Sepsis, jaundice |

*IgM, immunoglobulin M; Z, Zika; D, dengue; C, chikungunya; JUTH, Jos University Teaching Hospital; OLA, Our Lady of Apostles Hospital

†103 total infants born to IgM-positive mothers from May 2019–May 2021 had delivery and/or follow-up records in the study.

‡Transient weight and head circumference are defined as weight or head circumference <-2 Z-score for infant sex and age, adjusted for gestational age if preterm, at any infant visit from birth to 6 months, but normal at another visit. Transient can be low at birth and normal at follow-up, or normal at birth and low at follow-up, as indicated.

Supplementary Table S3. Symptoms associated with Zika, dengue, and chikungunya infection (Chembio IgM+) among pregnant women in Jos, Nigeria, April 2019-January 2022*

| | ZIKV/DENV/CHIKV negative or IgG+ only, no. (%)† | ZIKV IgM+, no. (%) | DENV IgM+, no. (%) | CHIKV IgM+, no. (%) | Multiple IgM+, no. (%) | Fisher's exact p value |
|---|---|---|---|---|---|---|
| Zika symptoms | | | | | | |
| Zika screening status‡ | | | | | | |
| Asymptomatic | 150 (18.6) | 4 (28.6) | 22 (36.1) | 24 (31.2) | 19 (38.8) | <.001 |
| Symptomatic | 655 (81.4) | 10 (71.4) | 39 (63.9) | 53 (68.8) | 30 (61.2) | |
| Reported Zika symptoms† | | | | | | |
| Asymptomatic | 107 (13.3) | 4 (28.6) | 15 (24.6) | 13 (16.9) | 10 (20.4) | .037 |
| Symptomatic | 698 (86.7) | 10 (71.4) | 46 (75.4) | 64 (83.1) | 39 (79.6) | |
| Fever | | | | | | |
| No | 503 (62.5) | 8 (57.1) | 34 (55.7) | 46 (59.7) | 21 (42.9) | .079 |
| Yes | 302 (37.5) | 6 (42.9) | 27 (44.3) | 31 (40.3) | 28 (57.1) | |
| Headache | | | | | | |
| No | 210 (26.1) | 6 (42.9) | 22 (36.1) | 21 (27.3) | 13 (26.5) | .306 |
| Yes | 595 (73.9) | 8 (57.1) | 39 (63.9) | 56 (72.7) | 36 (73.5) | |
| Arthralgia | | | | | | |
| No | 699 (86.8) | 12 (85.7) | 53 (86.9) | 65 (84.4) | 36 (75.0) | .240 |
| Yes | 106 (13.2) | 2 (14.3) | 8 (13.1) | 12 (15.6) | 12 (25.0) | |
| Myalgia | | | | | | |
| No | 726 (90.2) | 12 (85.7) | 50 (82.0) | 68 (88.3) | 42 (87.5) | .265 |
| Yes | 79 (9.8) | 2 (14.3) | 11 (18.0) | 9 (11.7) | 6 (12.5) | |
| Rash | | | | | | |
| No | 771 (95.8) | 14 (100) | 58 (95.1) | 72 (93.5) | 42 (87.5) | .120 |
| Yes | 34 (4.2) | 0 (0) | 3 (4.9) | 5 (6.5) | 6 (12.5) | |
| Conjunctivitis | | | | | | |
| No | 771 (95.9) | 14 (100) | 61 (100) | 72 (93.5) | 44 (91.7) | .147 |
| Yes | 33 (4.1) | 0 (0) | 0 (0) | 5 (6.5) | 4 (8.3) | |
| Additional symptoms | | | | | | |
| Fatigue | | | | | | |
| No | 567 (71.6) | 11 (78.6) | 41 (67.2) | 52 (68.4) | 21 (43.8) | .003 |
| Yes | 225 (28.4) | 3 (21.4) | 20 (32.8) | 24 (31.6) | 27 (56.3) | |
| Abdominal pain | | | | | | |
| No | 655 (83.2) | 10 (71.4) | 52 (85.3) | 62 (82.7) | 35 (72.9) | .280 |
| Yes | 132 (16.8) | 4 (28.6) | 9 (14.8) | 13 (17.3) | 13 (27.1) | |
| Nausea | | | | | | |
| No | 671 (85.0) | 13 (92.9) | 46 (75.4) | 64 (84.2) | 34 (72.3) | .059 |
| Yes | 118 (15.0 | 1 (7.1) | 15 (24.6) | 12 (15.8) | 13 (27.7) | |
| Vomiting | | | | | | |
| No | 668 (84.7) | 12 (92.3) | 45 (75.0) | 65 (85.5) | 33 (70.2) | .036 |
| Yes | 121 (15.3) | 1 (7.7) | 15 (25.0) | 11 (14.5) | 14 (29.8) | |
| Eye pain | | | | | | |
| No | 759 (96.1) | 14 (100) | 60 (98.4) | 73 (96.1) | 44 (93.6) | .772 |
| Yes | 31 (3.9) | 0 (0) | 1 (1.6) | 3 (4.0) | 3 (6.4) | |
| Joint swelling | | | | | | |
| No | 772 (98.1) | 14 (100) | 58 (96.7) | 76 (100) | 46 (100) | .558 |
| Yes | 15 (1.9) | 0 (0) | 2 (3.3) | 0 (0) | 0 (0) | |

*IgM, immunoglobulin M; ZIKV, Zika virus; DENV, dengue virus; CHIKV, chikungunya virus; IgG, immunoglobulin G.
†Column percentages shown.
‡Zika screening status was symptomatic if the participant reported any of the six Zika symptoms at the time of recruitment for the screening. Reported Zika symptoms are the symptoms reported when asked to provide symptom details in the questionnaire. Some participants were recruited for the screening as asymptomatic but reported one or more Zika symptoms in the questionnaire.
